# Validation of patient-reported outcome measures for postpartum depression, fatigue, maternal-infant bonding, and quality of life in Nigeria

**DOI:** 10.1101/2025.03.22.25323907

**Authors:** AI Abioye, TA Adeyemo, M Badmus, CF Chieme, H Adelabu, R Quao, T Lawanson, VO Adaramoye, OR Akinajo, M Balogun, A Banke-Thomas, OA Babah, E Igho-Osagie, EJ Mitchell, KF Walker, BB Afolabi, IVON-PP Trial Investigators

**Author notes:** Correspondence to: Professor Bosede B. Afolabi, Department of Obstetrics and Gynaecology, Faculty of Clinical Sciences, College of Medicine, University of Lagos, P.M.B 12003, Idi-Araba, Surulere, Lagos State, Nigeria. Funding: Bill & Melinda Gates Foundation, INV - 032911. Data described in the manuscript, code book, and analytic code will be made available upon reasonable request, and after obtaining approval from regulatory authorities of partner organizations.

## Abstract

**Background:** Postpartum anaemia significantly impacts maternal health globally. It is associated with depression, fatigue, lack of maternal-infant bonding, and poor quality of life in affected **mothers**. In this study we translated and validated patient-reported outcome measures (PROMs) for complications associated with postpartum anaemia in Nigerian women.

**Methods:** The Edinburgh Postnatal Depression Scale (EPDS), the Fatigue Severity Scale (FSS-5R), the Maternal-Infant Bonding Scale (MIBS), and the World Health Organization Quality of Life Brief Scale (WHOQOL-BREF) were translated in duplicate into Hausa, Nigerian Pidgin English (NPE) and Yoruba languages, and back-translated into English. We assessed face validity and internal consistency of the translated instruments during preparation for the IVON-PP trial (Intravenous ferric carboxymaltose versus oral ferrous sulphate for the treatment of moderate to severe postpartum anaemia in Nigerian women) among postpartum women (N=2,452). Logistic regression models evaluated the relationship of the PROMs to the odds of postpartum anaemia. We also assessed estimated intra-cluster correlation coefficients (ICC) for test-retest reliability among 335 women with repeated visits during the IVON-PP trial.

**Results:** The Cronbach’s alpha was good (>0.7) for the original English version of all PROMs, the FSS-5R in all languages, and the MIBS in Hausa and NPE. Test-retest reliability was moderate for EPDS (ICC=0.61), MIBS (ICC=0.60), and WHOQOL-BREF (ICC=0.60) in English, and for EPDS (ICC=0.77), FSS-5R (ICC=0.76), and WHOQOL-BREF (ICC=0.58) in Yoruba. Test-retest reliability was poor for the Hausa versions and could not be assessed for NPE, as there were too few participants (n=4). Postpartum anaemia was observed among 16.1% (395/2,452) of participants, and was significantly associated with the scores obtained with the instruments.

**Conclusion:** We translated and validated four postpartum anaemia PROMs in Nigeria, finding mixed reliability and predictive ability, making them valid alternatives to the English versions but we recommend re-evaluating the Hausa and NPE versions in larger studies.

## INTRODUCTION

Postpartum anaemia poses a significant burden of disease globally (1). When severe, postpartum anaemia doubles the risk of maternal mortality (2). About 80% of new mothers in low-income countries and as high as 50% of new mothers in high-income countries experience anaemia in the postpartum period(3). The condition results from untreated anaemia in the antenatal period or from significant obstetric haemorrhage(1,4,5). Postpartum anaemia disrupts maternal-infant bonding through its impact on fatigue which prevents adequate breastfeeding (1,6,7). Postpartum anaemia is also associated with an increased risk of infection(8,9), poor wound healing(10), slower maternal cognition(11), and depression(11,12), and these also have implications for maternal-infant bonding(11).

Several outcomes associated with postpartum anaemia e.g., fatigue, maternal bonding, and depression can only be captured in research using patient-reported outcome measures (PROMs). PROMs are questionnaires that collect information directly from patients about specific conditions, symptoms, quality of life, or functioning(13). Some of them include the Edinburgh Postnatal Depression Scale (EPDS)(14,15), the Fatigue Severity Scale (FSS-5R)(16,17), the World Health Organization’s Quality of Life Brief Scale (WHOQOL-BREF)(18,19), and the Mother-to-Infant Bonding Scale (MIBS)(20). These scales, although validated across several populations, have not been appropriately translated and validated for use in Nigerian native languages. The previous validation efforts in Nigeria have been restricted to EPDS in Yoruba(21,22) or among other clinical populations(23).

PROMs are usually developed and validated in specific populations, with the expectation that they would be reliablevalid, and, therefore, useful for ascertaining outcomes relevant to patients and other stakeholders across multiple countries and settings(24,25). PROMs are referred to as reliable if their responses tend to be consistent in respondents across their component items and when repeatedly administered. A PROM is regarded as valid if it truly measures the constructs it is intended to measure. When PROMs are adapted for use in a different population, it is best practice to evaluate their reliability and validity in that population, as different populations may interpret sentences and questions differently, or information may be lost when questionnaires are translated to other languages(26,27). Although English is the lingua franca, Nigeria is a multilingual society with more than 300 indigenous languages. The Hausa, Yoruba, and Igbo languages are the most widely spoken and are spoken by 32%, 17%, and 13%, respectively, as their primary languages (28). In addition, Nigerian Pidgin English (NPE), a patois formed from a combination of indigenous languages and English, is a common means of communication among Nigerians of different cultures or ethnicities in the country(29).

Trials conducted in countries where the primary language is not English may be incapable of capturing these important PROs because subjects may not understand the questions or may answer or interpret them differently from the populations in which they were originally validated. The objective of this study was to translate the EPDS, FSS-5R, MIBS, and WHOQOL-BREF scales into Nigerian languages (Hausa, Yoruba, and NPE) and evaluate the reliability and validity of the original English language versions among postpartum women in Nigeria. This work addresses a gap in the literature, as similar translations and validations of these scales in Nigerian languages have not been previously conducted. Hausa, Yoruba and NPE were selected as they were the major languages in the states the IVON-PP trial sites were located in. Adapting patient reported outcome measures into indigenous Nigerian languages can help clinical trial researchers, physicians and other stakeholders more robustly describe the burden of disease and the unmet patient needs of postpartum women in Nigeria.

## METHODS

### IVON-PP Trial

This validation study was conducted in preparation for an ongoing randomized controlled trial, the Intravenous Ferric Carboxymaltose Versus Oral Ferrous Sulphate For The Treatment Of Moderate To Severe Postpartum Anaemia In Nigerian Women (IVON-PP) Trial(30). The IVON-PP trial is an open-label trial assessing the clinical effectiveness, tolerability, and safety of intravenous ferric carboxymaltose versus oral ferrous sulphate among postpartum women in Nigeria with moderate to severe anaemia. The trial is being conducted at 20 primary, secondary, and tertiary health centers in Kano, Kwara, Lagos and Rivers states of Nigeria, selected for the presence of postnatal and immunization clinic services. Participants were enrolled into the trial 6-48 hours after birth and followed up for six months. The primary endpoint of the trial is the proportion of participants who remain anaemic at 6 weeks postpartum. The IVON-PP trial commenced in December 2022. It utilizes a hybrid effectiveness-implementation design to assess intervention effectiveness and collect contextual information for implementation (30).

Patient-reported secondary endpoints were assessed at 6 weeks and 6 months and include the prevalence of postpartum depression measured using the EPDS(15), fatigue measured using the FSS-5R(16,17), impaired maternal-infant bonding measured using the MIBS(20), and quality of life measured using the WHOQOL-BREF(18,19). We conducted a validation project as part of the cross-cultural adaptation of these PROMs for the IVON-PP trial. This project was based on two studies: 1) a cross-sectional survey and 2) a cohort study nested in the IVON-PP trial.

### Patient-reported outcome measures

#### Edinburgh postnatal depression scale (EPDS)

The EPDS is a validated instrument used to assess depression among postpartum women(31). The 10-item instrument asks participants about their feelings, mood, and thoughts about self-harm. Each item is rated on a Likert-type scale from 0 – 3, depending on whether and how often they have the adverse experience. The summary score is a sum of the individual ratings, with a range of possible values of 0 – 30. A score above 13 is used to determine the presence of depression(32).

#### Fatigue severity scale, 5-item, revised (FSS-5R)

The FSS-5R is a brief, specific, reliable, and valid measure for postpartum fatigue(33). Its five items ask participants whether and to what extent fatigue impacts their physical functioning. Each item is rated from 1 – 7, with a low score indicating that the statement is not very appropriate and a high score indicating agreement. A summary score is obtained as the mean score for all items. The range of possible values for the FSS-5R is, therefore, from 1 to 7. A threshold of >4 is used to determine the presence of fatigue(34).

#### Maternal infant bonding scale (MIBS)

The MIBS is validated for assessing mothers’ moods in the postpartum period. It also serves as a predictor for subsequent postpartum depression(20). It assesses maternal-infant bonding in four domains: impaired bonding, rejection and pathological anger, infant-focused anxiety, and incipient abuse. The scale comprises twenty-five items that assess concepts related to a mother’s interaction with the baby: whether they feel close to the baby, are joyful and pleased with motherhood, resent the baby, are anxious, or regret motherhood. Each item is rated on a six-point Likert scale (from 0, “never” to 5, “always”), with the scale of some items reversed. The MIBS score is calculated as a sum of the values for each item, and the total scores range from 0 to 125. A high score indicates worse mother-to-infant bonding. Specific thresholds exist for the domains: impaired bonding (>12), rejection and pathological anger (>13), infant-focused anxiety (>10), and incipient abuse (>3).

#### World Health Organization Quality of Life Brief Scale (WHOQOL-BREF)

The WHOQOL-BREF assesses quality of life(35). The scale has twenty-six items – two items that assess perceived quality of life and satisfaction with health, and twenty-four questions that assess four domains: physical, psychological, social relationships and environment. Each item was scored on a five-point Likert scale from “not at all” to “extremely” or similar phrasing, with scores ranging from 1 – 5(36). The summary score is a sum of the scores from each item and then transformed to a scale with a maximum score of 100. A threshold of <60 was used to determine the presence of impaired quality of life(36).

### Translation

All PROMs were translated into Hausa, Yoruba and Nigerian Pidgin English. Instrument translation and cultural adaptation of the PROMs followed guidelines outlined in Beaton et al. (2000) and by the International Society of Pharmacoeconomics and Outcomes Research (26,27). For each language, two individuals, a certified translator, and an expert translator, translated each instrument; both translations were back-translated to English by a third individual to compare and reconcile the differences. A team of the investigators and translators reconciled discrepancies, leading to a single translated version of each instrument. The instrument was pre-tested by 10 health workers who administered them to one randomly selected patient each at selected study sites, and further translation and understanding discrepancies were resolved. The final translated instruments were then validated.

### Cross-sectional survey

#### Study population and study design

The PROMs were administered in a cross-sectional survey conducted at IVON-PP sites in December 2022, before the commencement of the trial. 2,452 eligible women were screened, gave their consent and were enrolled into the study. Participants were eligible if they were 15 – 49 years’ old, attending the postpartum clinic at approximately 6 weeks’ postpartum, and fluent in the selected or most spoken language for the region. There were no restrictions on participants’ health status. Participants completed the four PROMs in only one language, selected as the language most spoken in the state, or pre-selected by the researchers. Thus, participants in Lagos responded to English translations, those in Kano, Kwara and Rivers states responded to questionnaires in Hausa, Yoruba and Nigerian Pidgin English respectively. A test-retest reliability assessment was done to evaluate the consistency of the responses over time. Participants provided sociodemographic information, including age, parity, marital status and level of education. The surveys were interviewer-administered electronically using REDCap. All participants had blood samples collected for point of care hemoglobin testing at the visit with the use of Hemocue HB 801 meter.

#### Sample size

There are no standard guidelines for estimating the minimum sample size for instrument validation studies. The minimum sample size for each language in this study was estimated based on a rule of thumb: the number of questions in the longest instrument, multiplied by a factor of 20(37). Of the four PROMs, the WHOQOL-BREF was the longest, with 30 questions(35). Therefore, the minimum sample size was 600 for each language, and the total minimum sample size was 2,400.

#### Descriptive summaries of PROMs

Each PROM in each language was summarized using the median, lower and upper quartiles, minimum and maximum, and the proportion meeting clinically relevant thresholds were estimate.

#### Content validity

Content validity was assessed by the investigators by reviewing the items of the instruments based on their expert opinion to determine whether the instruments were likely to truly assess the targeted concepts they were supposed to assess.

#### Face validity

Research nurses asked a subset of participants about the questionnaires’ comprehensibility, simplicity, length, and perceived relevance to the intended objective using a Likert-type scale, and favorable responses (such as very easy or easy) noted. Every 12th participant selected automatically through REDCap (minimum of 50 per language) was included in the face validity exercises.

#### Internal consistency

The scale reliability coefficient or Cronbach’s α was calculated to assess the internal consistency for each PROM in each language. Cronbach’s α of ≥0.7 and above was considered satisfactory (38).

#### Prediction of postpartum anaemia

The extent to which the PROMs distinguish someone with postpartum anaemia from a person without was evaluated in logistic regression models and odds ratios (with 95% confidence intervals) obtained. Postpartum anaemia was defined as hemoglobin <11 g/dL. Continuous and dichotomous values of the PROMs were considered in separate models.

### Test-retest or cohort study

A subset of participants in the IVON-PP trial were invited to retake the PROMs in their preferred languages within four weeks of the primary endpoint assessment which occurred at week six visit. Test-retest reliability was calculated by estimating intra-class correlation coefficients (ICC) which assessed the extent to which participants’ PROM scores agreed at the two time-points ≤4 weeks apart. The ICC model was based on a two-way mixed effects model for a single measurement rather than the mean of multiple measurements(39,40). The ICC is interpreted thus: <0.5 poor reliability, 0.5 – <0.75 moderate reliability, 0.75 – <0.9 good reliability and >0.9 excellent reliability(39). While the language for the PROMs was assigned to the different states based on the predominant language spoken there, participants chose their preferred language for the test-retest assessment.

### Ethical considerations

For the validation study, verbal informed consent was obtained after the study procedure was explained to participants, and participants’ personal information or identification were not collected. Written consent was obtained for the IVON-PP trial and verbal consent was obtained for the extra visit required for the test-retest study. Institutional ethical clearance for the validation study and the test-retest study as components of the IVON-PP trial was obtained from the National Health Research Ethics Committee, Nigeria (NHREC/01/01/2007-07/09/2022), the institutional review boards of the participating teaching hospitals and the States’ health ethics boards. Approval for the IVON-PP trial was also obtained from the National Agency for Food and Drug Administration and Control. The trial is registered in the International Standard Randomised Controlled Trial registry (ISRCTN51426226).

#### Data Analysis

Statistical analyses were conducted using the *Tableone*, *ltm*, and *irr* packages in RStudio 1.0.153(41–44). No imputation was done for missing items.

## RESULTS

2,452 trial participants completed PROMs in their preferred or designated language. 207 participants were selected to conduct face validation of the PROMs, while 335 participants retook their PROMs for the test-retest reliability assessment **(Table 1)**. The mean age of participants was 30 years (SD: 6). Sixty-five (65%) percent of them were in their first or second pregnancies (65%). While the language for the PROMs was pre-selected for the validation study according to the state, participants chose their preferred language for the test-retest study: English (50%), Hausa (23%), Yoruba (25%) and NPE (1%).

**Table 1.** Participant characteristics.

| Characteristics | Validation study,<br>N=2452 | Face validation sub-study,<br>N=207 | Test-retest, N=335 |
| --- | --- | --- | --- |
| State |  |  |  |
| Kano | 606 (25%) | 51 (25%) | 91 (27%) |
| Kwara | 625 (26%) | 51 (25%) | 123 (37%) |
| Lagos | 618 (25%) | 53 (26%) | 63 (19%) |
| Rivers | 603 (25%) | 52 (25%) | 58 (17%) |
| Language |  |  |  |
| English | 606 (25%) | 51 (25%) | 169 (50%) |
| Hausa | 625 (26%) | 51 (25%) | 78 (23%) |
| Nigerian Pidgin English | 618 (25%) | 53 (26%) | 4 (1%) |
| Yoruba | 603 (25%) | 52 (25%) | 83 (25%) |
| Age, y |  |  |  |
| Median (IQR) | 29 (25, 33) | 30 (25, 34) | 28 (24, 32) |
| <20 | 38 (2%) | 4 (2%) | 14 (5%) |
| 20 – <30 | 1202 (49%) | 97 (47%) | 171 (51%) |
| 30 – <40 | 1105 (45%) | 95 (46%) | 141 (42%) |
| ≥40 | 107 (4%) | 11 (5%) | 8 (2%) |
| Parity |  |  |  |
| 1 | 823 (34%) | 66 (32%) | 129 (39%) |
| 2 | 750 (31%) | 66 (32%) | 80 (24%) |
| 3 | 439 (18%) | 38 (18%) | 46 (14%) |
| 4+ | 440 (18%) | 37 (18%) | 79 (24%) |
| Marital status |  |  |  |
| Married/cohabiting | 2371 (97%) | 200 (97%) | 326 (97%) |
| Single, never married | 63 (3%) | 4 (2%) | 7 (2%) |
| Others | 18 (<1%) | 3 (1%) | 2 (1%) |
| Highest level of education |  |  |  |
| No formal education | 49 (2%) | 8 (4%) | 9 (3%) |
| Completed primary | 145 (6%) | 16 (8%) | 31 (9%) |
| Completed secondary | 1078 (44%) | 83 (40%) | 146 (44%) |
| Completed tertiary | 1121 (46%) | 98 (47%) | 127 (38%) |
| Postgraduate | 59 (2%) | 2 (1%) | 20 (6%) |

\*Percentages may not add up to 100% due to rounding.
\*\*The following variables had missing observations in the reliability cohort: parity (n=1), level of education (n=2)

**Figure 1.**
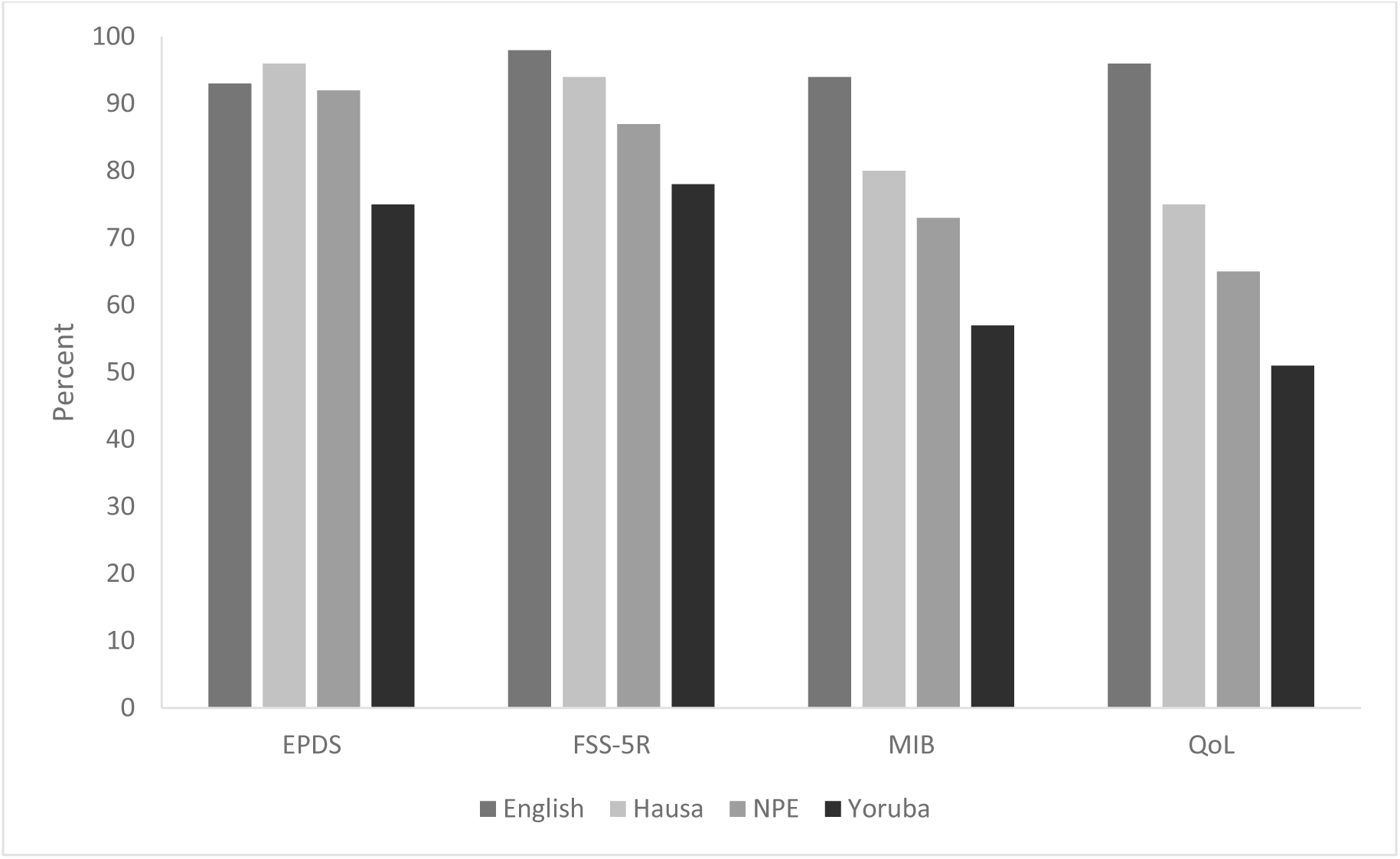
Face validity of PROMs

The median scores for all PROMs varied across the different languages in which they were administered (**Table 2**). Notably, PROMs completed in Nigerian Pidgin English (NPE) had the highest proportions of the measured outcomes, while those completed in Hausa had the lowest proportions. For example, the proportion of depression among participants that filled the EPDS in NPE and Hausa, respectively, were 28% and 3%, while the proportion of patients experiencing fatigue in both groups were 28% and 5%, respectively.

**Table 2.** Descriptive summaries of the measures in validation cohort (N=2452)

|  | English | Hausa | Nigerian Pidgin English | Yoruba |
| --- | --- | --- | --- | --- |
| EPDS |  |  |  |  |
| Min, Max | 0, 22 | 0, 22 | 0, 25 | 0, 24 |
| Median (IQR) | 8 (4, 11) | 2 (0, 6) | 12 (8, 14) | 8 (5, 10) |
| Depressed, >13 | 82 (13%) | 18 (3%) | 170 (28%) | 52 (8%) |
| FSS-5R |  |  |  |  |
| Min, Max | 1, 7 | 1, 7 | 1, 7 | 1, 7 |
| Median (IQR) | 2 (2, 3) | 1 (1, 2) | 3 (1, 4) | 2 (1, 2) |
| Fatigued, ≥4 | 95 (15%) | 32 (5%) | 166 (28%) | 50 (8%) |
| MIB score |  |  |  |  |
| Min, Max | 0, 66 | 0, 65 | 0, 94 | 0, 114 |
| Median (IQR) | 6 (2, 10) | 13 (6, 23) | 18 (10, 28) | 6 (3, 10) |
| Impaired bonding score |  |  |  |  |
| Min, Max | 0, 33 | 0, 31 | 0, 44 | 0, 54 |
| Median (IQR) | 3 (2, 6) | 10 (6, 14) | 8 (4, 13) | 4 (2, 7) |
| Impaired bonding, >12 | 22 (4%) | 213 (35%) | 167 (28%) | 19 (3%) |
| Rejection and anger score |  |  |  |  |
| Min, Max | 0, 20 | 0, 19 | 0, 27 | 0, 35 |
| Median (IQR) | 0 (0, 1) | 1 (0, 5) | 5 (2, 8) | 0 (0, 1) |
| High rejection and anger score, >12 | 7 (1%) | 15 (3%) | 24 (4%) | 2 (0.3%) |
| Infant focus anxiety |  |  |  |  |
| Min, Max | 0, 12 | 0, 11 | 0, 13 | 0, 15 |
| Median (IQR) | 0 (0, 2) | 1 (0, 3) | 5 (3, 6) | 0 (0, 2) |
| Anxious about infant, >12 | 0 (0%) | 0 (0%) | 2 (0.3%) | 1 (0.2%) |
| Incipient abuse score |  |  |  |  |
| Min, Max | 0, 9 | 0, 7 | 0, 10 | 0, 10 |
| Median (IQR) | 0 (0, 0) | 0 (0, 2) | 0 (0, 1) | 0 (0, 0) |
| High incipient abuse score, >12 | 0 (0%) | 0 (0%) | 0 (0%) | 1 (0.2%) |
| WHOQOL |  |  |  |  |
| Min, Max | 25, 100 | 19, 100 | 19, 100 | 19, 100 |
| Median (IQR) | 63 (57, 69) | 75 (69, 82) | 63 (57, 69) | 63 (50, 69) |
| Impaired quality of life, <60 | 224 (36%) | 38 (6%) | 240 (40%) | 289 (46%) |

### Face validity

Broadly, participants completed each of the interviewer-administered questionnaires at an average of <5 minutes (**Supplementary table 1)** and they regarded the questionnaires as easy to understand, simple to respond to and not too lengthy (**Supplementary table 3)**. The EPDS and FSS-5R were scored as “easy or very easy to understand”, as having “simple or very simple questions”, as being a “short or very short questionnaire” and being more likely to identify the outcome measured by the PROM by more Hausa respondents compared with other languages. English responding participants were more likely to regard the MIB and QOL instruments as easy to understand, simple to respond to and not too lengthy. Overall, lower proportions of Yoruba responding participants regarded the PROMs as easy to understand, short, easy to answer or likely to elicit the outcomes they were designed for. Respondents across all language groups were less likely to indicate that the questionnaires were short or very short; ranging from 10% (EPDS, Yoruba) to 88% (EPDS and FSS-5R, Hausa). Overall, most participants noted that the questionnaires measured the outcomes they were designed for. The proportion of respondents who indicated yes to that question was greater than 90% for all PROMs and for all languages apart from the Yoruba respondents.

### Reliability

#### Internal consistency

The Cronbach’s alpha was good (>0.7) for the original English version of all PROMs **(Table 3).** For the translated versions, Cronbach’s α was good for the FSS-5R in all languages (α=0.95 for Hausa, α=0.92 for NPE, and α=0.90 for Yoruba), and the MIBS in Hausa (α=0.92) and NPE (α=0.88). Cronbach’s α was poor for EPDS in NPE and Yoruba, MIB in Yoruba, and WHOQOL in Hausa and Yoruba.

**Table 3.** Internal consistency and test-retest reliability.

|  | English | Hausa | Nigerian Pidgin English | Yoruba |
| --- | --- | --- | --- | --- |
| Internal consistency (N=2,452)<sup>1</sup> |  |  |  |  |
| EPDS | 0.74 (0.72, 0.77) | 0.69 (0.63, 0.73) | 0.57 (0.52, 0.62) | 0.57 (0.52, 0.61) |
| FSS-5R | 0.89 (0.87, 0.91) | 0.95 (0.94, 0.96) | 0.92 (0.90, 0.93) | 0.90 (0.87, 0.92) |
| MIB | 0.80 (0.75, 0.84) | 0.92 (0.91, 0.93) | 0.88 (0.87, 0.89) | 0.68 (0.61, 0.75) |
| Impaired bonding | 0.60 (0.52, 0.67) | 0.81 (0.78, 0.83) | 0.76 (0.73, 0.78) | 0.50 (0.39, 0.60) |
| Rejection and anger | 0.65 (0.55, 0.73) | 0.83 (0.81, 0.85) | 0.73 (0.70, 0.76) | 0.55 (0.34, 0.53) |
| Infant-focused anxiety | 0.46 (0.38, 0.52) | 0.41 (0.33, 0.48) | 0.33 (0.24, 0.41) | 0.26 (0.17, 0.34) |
| Incipient abuse | 0.41 (0.18, 0.72) | 0.88 (0.84, 0.92) | 0.77 (0.70, 0.82) | 0.54 (0.04, 0.78) |
| WHOQOL-BREF | 0.79 (0.76, 0.81) | 0.53 (0.42, 0.61) | 0.67 (0.62, 0.70) | 0.60 (0.55, 0.64) |
| Test-retest reliability (N=335)<sup>2</sup> |  |  |  |  |
| EPDS | 0.61 (0.51, 0.70) | 0.21 (-0.02, 0.41) |  | 0.77 (0.67, 0.85) |
| FSS-5R | 0.30 (0.16, 0.43) | -0.03 (-0.25, 0.19) |  | 0.76 (0.64, 0.84) |
| MIB | 0.60 (0.49, 0.70) | 0.03 (-0.20, 0.25) |  | 0.48 (0.29, 0.64) |
| Impaired bonding | 0.52 (0.40, 0.63) | 0.11 (-0.12, 0.33) |  | 0.55 (0.37, 0.69) |
| Rejection and anger | 0.48 (0.35, 0.60) | 0.11 (-0.12, 0.33) |  | 0.22 (-0.01, 0.42) |
| Infant-focused anxiety | 0.70 (0.61, 0.77) | 0.30 (0.08, 0.50) |  | 0.27 (0.05, 0.47) |
| Incipient abuse | 0 (-0.16, 0.16) | 0 (-0.22, 0.22) |  | 0 (-0.23, 0.23) |
| WHOQOL-BREF | 0.60 (0.49, 0.69) | 0.46 (0.26, 0.62) |  | 0.58 (0.41, 0.71) |

1. Cronbach's alphas are reported for internal consistency.
2. Intraclass correlations are reported for test-retest reliability

#### Test-retest reliability

Intraclass correlations (ICC) were ‘good’ for the Yoruba translations of the EPDS (0.77) and FSS-5R (0.76), while the ICC scores for the English versions of the WHOQOL-BREF and MIB total score (both 0.6) and the Yoruba translation of the WHOQOL-BREF (0.58) were moderate. The remaining ICC scores were observed to be poor (<0.5) for the English and Yoruba versions. The test-retest reliability (Table 2) was poor for all the PROMs in the Hausa group (all ICC < 0.5). The extra visit for the test-retest reliability was conducted on a median (IQR) of 28 (25, 29) days before the six-week postpartum visit. The median number of days was similar for English, Hausa, and Yoruba, though there was less variation in the Hausa group (**Supplementary Figure 1**). Test-retest reliability data was dropped in the NPE group as there were too few people (N=4).

### Prediction

The prevalence of postpartum anaemia among participants in the validation study was 16.1% (395/2,452). **Table 4** presents the results of the prediction of postpartum anaemia assessed using regression analysis. EPDS >13 indicative of depression was significantly related to postpartum anaemia in the English (OR=2.25; 95%CI: 1.34, 3.71), and Hausa language (OR=8.05; 95%CI: 3.04, 21.33) versions of the scale. In addition, the continuous form of the EPDS was significantly related to anaemia in the NPE version (OR=1.05; 95%CI: 1.01, 1.11). FSS-5R >4 was significantly related to anaemia in the Hausa (OR=5.75; 95%CI: 2.66, 12.2) and NPE (OR=1.74; 95%CI: 1.19, 2.68) versions. There were significant relations of the rejection and pathological anger domain in the Hausa (OR=11.65; 95%CI: 3.25, 46.54), and the infant-focused anxiety domain in the NPE version (OR= 1.80; 95%CI:1.08, 3.14). WHOQOL-BREF <60% was significantly related to the risk of anaemia in the Hausa (OR=3.27; 95%CI: 1.49, 6.75) and NPE (OR=2.17; 95%CI: 1.43, 3.31) versions. The continuous form of the WHOQOL-BREF in the Yoruba version was also significantly related to anaemia (OR= 0.98; 95%CI: 0.96, 0.99).

**Table 4.** Prediction – relationship to postpartum anaemia in validation survey (N=2452)

|  | English<br>OR (95%CI) | Hausa<br>OR (95%CI) | Nigerian Pidgin English<br>OR (95%CI) | Yoruba<br>OR (95%CI) |
| --- | --- | --- | --- | --- |
| EPDS |  |  |  |  |
| Continuous | 1.10 (1.05, 1.14) | 1.08 (1.03, 1.13) | 1.05 (1.01, 1.11) | 0.99 (0.94, 1.06) |
| >13 | 2.25 (1.34, 3.71) | 8.05 (3.04, 21.33) | 1.24 (0.79, 1.93) | 0.83 (0.31, 1.87) |
| FSS-5R |  |  |  |  |
| Continuous | 0.97 (0.83, 1.14) | 1.53 (1.26, 1.86) | 1.18 (1.04, 1.34) | 1.05 (0.86, 1.26) |
| >4 | 1.01 (0.58, 1.72) | 5.75 (2.66, 12.15) | 1.74 (1.19, 2.68) | 1.70 (0.77, 3.41) |
| Impaired bonding |  |  |  |  |
| Continuous | 1.00 (0.6, 1.05) | 1.04 (0.99, 1.09) | 1.03 (0.99, 1.07) | 1.01 (0.96, 1.05) |
| >12 | 1.45 (0.51, 3.61) | 1.22 (0.73, 2.00) | 1.41 (0.90, 2.18) | 1.30 (0.30, 4.04) |
| Rejection and anger |  |  |  |  |
| Continuous | 1.00 (0.91, 1.05) | 1.06 (0.99, 1.13) | 1.07 (1.02, 1.13) | 0.94 (0.82, 1.05) |
| >13 | NR | 11.65 (3.25, 46.54) | 1.51 (0.48, 4.00) | NR |
| Infant focused anxiety |  |  |  |  |
| Continuous | 1.04 (0.95, 1.15) | 1.09 (0.98, 1.20) | 1.13 (1.03, 1.23) | 1.04 (0.92, 1.17) |
| >10 | 1.28 (0.73, 2.15) | 1.71 (0.91, 3.07) | 1.80 (1.08, 3.14) | 1.06 (0.49, 2.08) |
| Incipient abuse |  |  |  |  |
| Continuous | 1.00 (0.71, 1.29) | 1.01 (0.84, 1.21) | 1.07 (0.91, 1.23) | 0.89 (0.41, 1.27) |
| >3 | NR | NR | NR | NR |
| Quality of Life |  |  |  |  |
| Continuous | 1.00 (0.98, 1.01) | 0.97 (0.95, 0.99) | 0.97 (0.95, 0.99) | 0.98 (0.96, 0.99) |
| <60% | 1.12 (0.75, 1.67) | 3.27 (1.49, 6.75) | 2.17 (1.43, 3.31) | 1.39 (0.87, 2.21) |

Values are RR (95%CI). Shouldn't we mention that logistic regression was used here?

## DISCUSSION

To the best of our knowledge, this is the first time these PRO instruments have been translated into these languages and comprehensively validated in Nigerian women in the postpartum period. In our study, we observed that PROMs completed in Nigerian Pidgin English had the highest proportions of the outcomes assessed by the PROMs, while patients completing the PROMs in Hausa had the lowest proportions of respective outcomes. It is impossible to conclude that patients in Rivers State where the NPE translation was administered were more depressed, fatigued or had lower quality of life than patients in Kano state where Hausa translations were administered. Results may reflect cultural differences in how women in the different Nigerian states respond to questions about their health or about the severity of health states. We note that this is plausible because the regions where the PROMs were administered are culturally and tribally different(45). Observed differences in proportions of measured outcomes may also have been due to differences in translation/perception of the items in the instruments. Future studies could focus on administering different language versions of the PROMs to populations of women who are multilingual.

Reliability results appeared to be mixed in our study. For example, Cronbach’s alpha values for all English language versions of the assessed PROMs suggest that they are internally consistent in English speaking Nigerians, but the ICC estimates demonstrated that only the WHOQOL-BREF and MIB English versions were reproducible. Reliability using Cronbach’s alphas was also demonstrated in all three Nigerian translations of the FSS-5R, implying that this study’s FSS-5R translations may be suitable for future studies in Nigeria. However, ICC estimations demonstrated that only the Yoruba translation of the FSS-5R was reproducible. Cronbach’s alphas for all translations of the WHOQOL-BREF indicated poor internal consistency, but the Yoruba translation’s ICC scores were determined to show moderate reproducibility. The lack of reproducibility may suggest that the PROMs were assessed too far apart. Although there are no standard guidelines, a duration of two days to two weeks has been recommended by many investigators(46).

The prevalence of postpartum anaemia in this study was 19%, consistent with results reported from another cross-sectional study (20%) in Lagos(47), but much lower than the 48% reported from an observational study in Enugu, Nigeria(48). Postpartum anaemia is associated with significant consequences for the woman, her offspring, and the larger society(1,6,7). As observed in this study, depression, fatigue, impaired maternal-infant bonding, and poor quality of life are important patient-relevant outcomes for women with postpartum anaemia. We also found probable postpartum depression among 13%, fatigue among 15%, and impaired quality of life among 36% of participants in this study based on the English language version of the instruments. Several researchers have investigated the association of these patient-reported outcomes with maternal anaemia or as endpoints in iron supplementation trials: postpartum depression(14,49–51), fatigue(14,50,52,53), and quality of life(51). These studies had mixed results, ranging from significant positive response(49,51,53) or null effect(14,50,52). In this study, we found significant relations between anaemia and poor quality of life when using the translated versions of the WHOQOL-BREF. The other PROMs were not associated with anaemia, even in their English-language versions, at the prespecified clinically relevant thresholds. The challenge may be in the severity or type of anaemia outcome itself. The absence of significant findings in the regression analyses may also be related to limited statistical power. This is illustrated by the significant findings in some of the analyses using the continuous form of PROMs.

This study contributes significantly to the body of psychometric knowledge. As noted, it is likely the first to translate all four PROMs into Nigerian languages of Yoruba, Hausa, and Nigerian pidgin with demonstrated reliability and validity. The study also has some noteworthy limitations. We did not simultaneously evaluate any of the participants using both the English language version of the instrument and a translated version. We were thus unable to assess the agreement, sensitivity, and specificity of the translated instruments in relation to the original English language version. While we could have done this had we specifically enrolled multilingual speakers, we note that even the English language version of these instruments requires validation in a Nigerian population, given potential differences in how English is spoken or understood. Given the geographic and sociodemographic diversity, we are fairly confident that our results may be generalizable to all postpartum Nigerian women. Third, given that participants responded to the four instruments at each visit, it is plausible that the effort required may have influenced some of the responses. Unfortunately, we are unable to isolate and address this *post-hoc.* Finally, the number of days between the visits for the test-retest assessment did not sufficiently vary, making it impossible to explore whether the test-retest reliability measure would have changed if the time between the visits were shorter.

## CONCLUSION

We found mixed results on the reliability of the instruments and ability of the PROMs to predict anaemia outcomes. Future iterations may leverage detailed item-response analysis to guide improvements to the PROMs. Therefore, we conclude that the translated PROMs are acceptable as alternatives to the English version, though future improvements are necessary.

## Data Availability

All data produced in the present study are available upon reasonable request to the authors

## SUPPLEMENT

**S. Figure 1.**
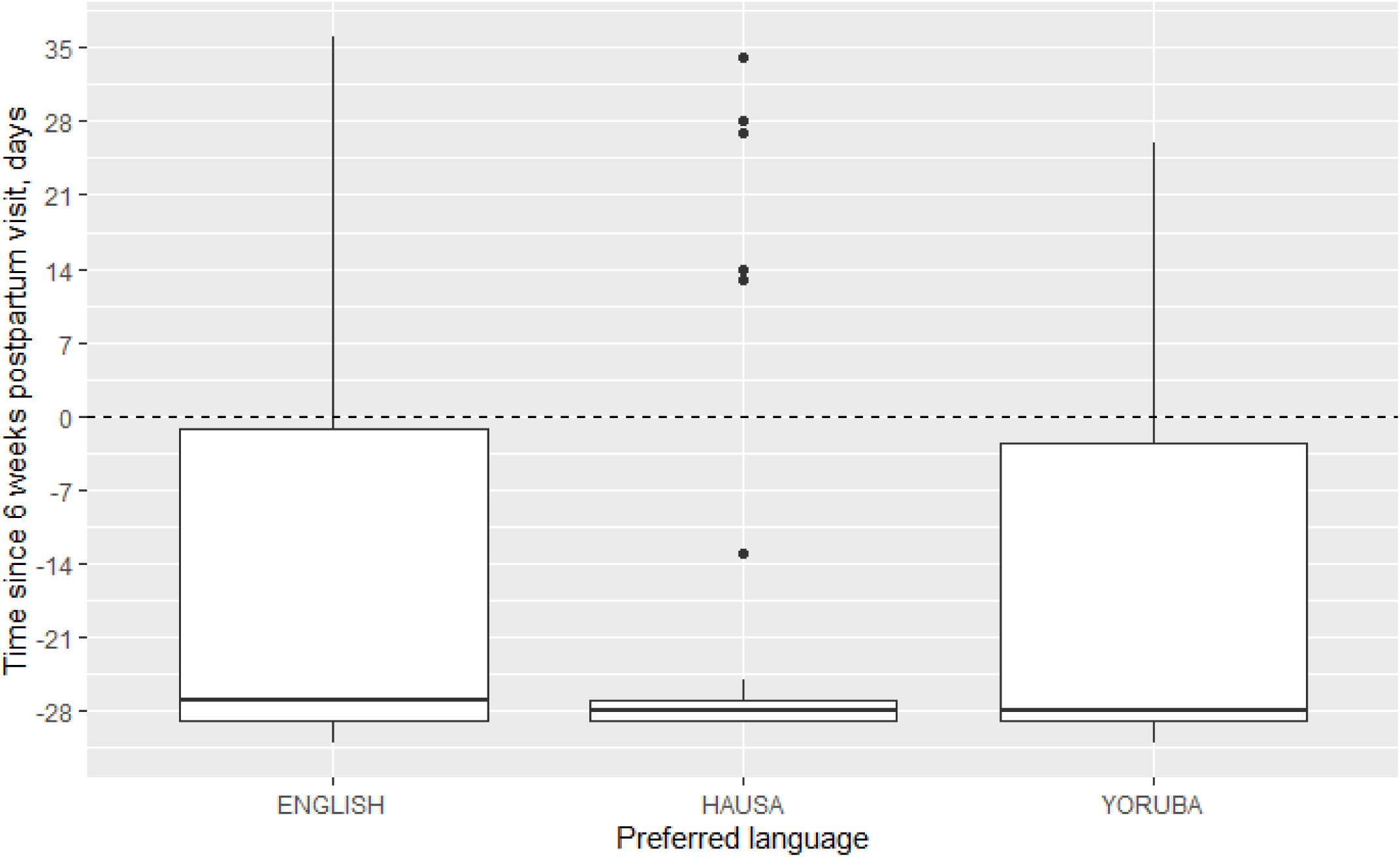
The difference in days between the assessment of the reliability measures

**S. Table 1.** Time spent in minutes in validation cohort (N=2452)

| Instrument | English,<br>Mean (SD) | Hausa,<br>Mean (SD) | Nigerian Pidgin English,<br>Mean (SD) | Yoruba,<br>Mean (SD) |
| --- | --- | --- | --- | --- |
| EPDS | 1.2 (0.7) | 0.8 (0.4) | 1.2 (1.3) | 1.1 (0.7) |
| Fatigue | 0.9 (1.0) | 0.4 (0.4) | 0.9 (1.1) | 0.6 (0.7) |
| MIB | 3.6 (6.7) | 1.5 (0.9) | 2.9 (1.8) | 1.9 (0.9) |
| QoL | 4.0 (2.1) | 2.1 (1.3) | 4.1 (3.1) | 4.0 (2.9) |

**S. Table 2.** Face Validity (N=207)

| Instrument |  | English, % | Hausa, % | Nigerian Pidgin English, % | Yoruba, % |
| --- | --- | --- | --- | --- | --- |
| EPDS | How easy was it for you to understand the question? | 47 (89%) | 47 (92%) | 45 (87%) | 29 (57%) |
|  | How simple was it for you to answer the question? | 42 (79%) | 49 (96%) | 42 (81%) | 25 (49%) |
|  | How lengthy do you consider this questionnaire? | 31 (59%) | 45 (88%) | 33 (64%) | 8 (16%) |
|  | Do you think the questions asked will make it possible to know if you have depression? | 53 (100%) | 51 (100%) | 48 (92%) | 38 (75%) |
|  | Face validity | 49 (92.5%) | 49 (96.1%) | 48 (92.3%) | 26 (56.9%) |
| FSS-5R | How easy was it for you to understand the question? | 50 (94%) | 48 (94%) | 44 (85%) | 39 (77%) |
|  | How simple was it for you to answer the question? | 45 (85%) | 48 (94%) | 40 (77%) | 37 (73%) |
|  | How lengthy do you consider this questionnaire? | 35 (66%) | 45 (88%) | 28 (54%) | 18 (35%) |
|  | Do you think the questions asked will make it possible to know if you have fatigue? | 50 (94%) | 46 (90%) | 41 (79%) | 32 (63%) |
|  | Face validity | 52 (98%) | 48 (94%) | 45 (87%) | 40 (78%) |
| MIB | How easy was it for you to understand the question? | 50 (94%) | 40 (78%) | 31 (60%) | 32 (63%) |
|  | How simple was it for you to answer the question? | 42 (79%) | 35 (79%) | 32 (62%) | 25 (49%) |
|  | How lengthy do you consider this questionnaire? | 22 (42%) | 20 (39%) | 10 (19%) | 5 (10%) |
|  | Do you think the questions asked will make it possible to determine how well you are bonding or establishing a good mother-infant relationship? | 52 (98%) | 51 (100%) | 50 (96%) | 39 (77%) |
|  | Face validity | 50 (94%) | 41 (80%) | 38 (73%) | 29 (57%) |
| QoL | How easy was it for you to understand the question? | 47 (89%) | 37 (73%) | 32 (62%) | 29 (57%) |
|  | How simple was it for you to answer the question? | 43 (81%) | 36 (71%) | 29 (56%) | 22 (43%) |
|  | How lengthy do you consider this questionnaire? | 31 (59%) | 23 (45%) | 6 (12%) | 11 (22%) |
|  | Do you think the questions asked will make it possible to assess your quality of life? | 53 (100%) | 51 (100%) | 49 (94%) | 35 (69%) |
|  | Face validity | 51 (96%) | 38 (75%) | 34 (65%) | 26 (51%) |

% refers to the proportion who respond "Very easy" or "Easy", "Simple" or "very simple", "short or very short", or "yes" as appropriate

**S. Table 3.** Summary of findings.

| Language | Instrument | Internally consistent | Test-retest reliable | Discriminant validity | Discrepant items | Face validity |
| --- | --- | --- | --- | --- | --- | --- |
| Hausa | EPDS | Good | No | Good | None | Yes |
|  | FSS-5R | Good | No | Good | None | Yes |
|  | MIBS | Good | No | Fair | None | Yes |
|  | WHOQOL-BREF | No | No | Good | None | Yes |
| Yoruba | EPDS | No | Yes | Poor | None | Yes |
|  | FSS-5R | Good | Yes | Poor | None | Yes |
|  | MIBS | Good | No | Poor | None | Yes |
|  | WHOQOL-BREF | No | Yes | Good | None | Yes |
| NPE | EPDS | No | No | Good | Yes | Yes |
|  | FSS-5R | Good | No | Good | None | Yes |
|  | MIBS | Good | No | Fair | None | Yes |
|  | WHOQOL-BREF | Good | No | Good | None | Yes |

**S. Table 4.**
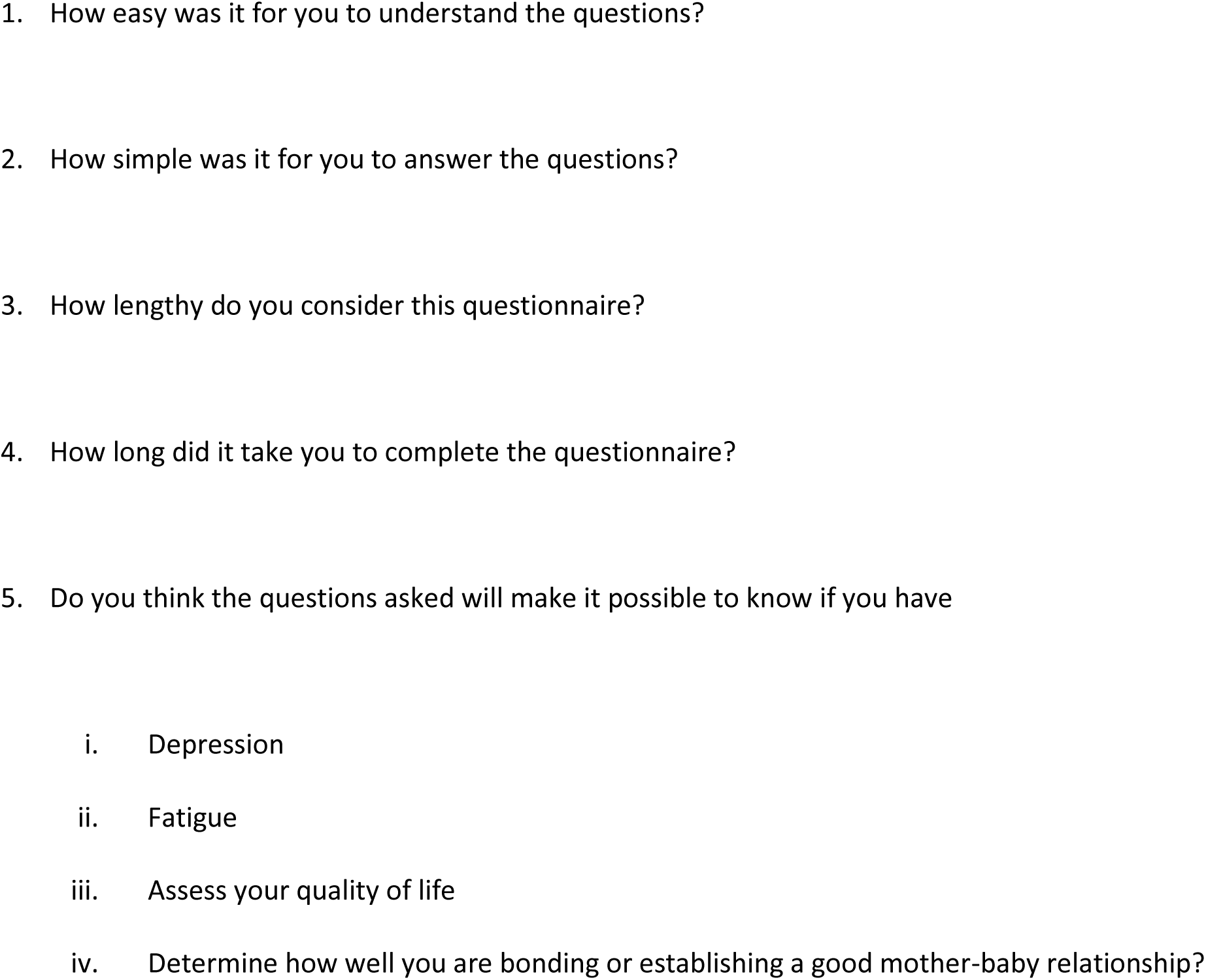
Questions for face validation of questionnaires.

